# Evidence for Impaired Homeostatic Regulation of Plasticity after Spinal Cord Injury

**DOI:** 10.64898/2026.03.24.26349041

**Authors:** Nahian S Chowdhury, Donovan Cheng, Stevan Nikolin, Yann Quidé, Negin Hesam-Shariati, Sylvia M Gustin

## Abstract

**Background:** Spinal cord injury (SCI) is associated with widespread reorganisation of cortical sensorimotor circuits. Persistent complications such as spasticity and neuropathic pain suggest that homeostatic plasticity, which normally helps stabilise and constrain activity-dependent changes in sensorimotor circuits, may be disrupted after SCI. Homeostatic plasticity can be probed using repeated blocks of transcranial direct current stimulation (tDCS); in healthy individuals, two closely spaced excitatory blocks typically leads to an inhibitory response, reflected as a reduction in corticomotor excitability.

**Objective:** To determine whether individuals with SCI show reduced homeostatic suppression of corticospinal excitability in response to repeated anodal tDCS, compared with healthy controls.

**Methods:** Twenty adults with thoracic or below SCI and 20 healthy controls completed three counterbalanced sessions. Each session comprised two 10-minute blocks of 2 mA tDCS separated by 5 minutes, with the second block always being anodal tDCS over left primary motor cortex. The first block was either anodal, cathodal, or sham tDCS, yielding 3 condition types: anodal-anodal, cathodal-anodal, and sham-anodal. To assess corticomotor excitability, transcranial magnetic stimulation-evoked motor evoked potentials (MEPs) were elicited at baseline, after priming, and every 5 minutes for 60 minutes after the second block. The primary outcome was percent change in MEP amplitude from baseline.

**Results:** In the anodal-anodal condition, the SCI group showed greater facilitation than controls over 0-30 minutes (estimate = 83.09, 95% CI 49.75 to 116.43, p < 0.001), suggestive of a weaker homeostatic response. The cathodal-anodal condition led to a significant overall facilitatory effect with no between-group difference, while the sham-anodal condition showed no change in MEP amplitude relative to baseline. Within the SCI group, exploratory subgroup analysis suggests that those with neuropathic pain and a traumatic injury showed greater facilitation in the anodal-anodal condition than those without these features, indicative of a weaker homeostatic response.

**Conclusions:** SCI is associated with impairment in the homeostatic regulation of corticomotor excitability following repeated excitatory brain stimulation. Disrupted plasticity stabilisation may be relevant to persistent symptoms such as neuropathic pain.

## Introduction

Spinal cord injury (SCI) causes disruption in sensorimotor pathways and widespread reorganization in cortical circuits [1]. In both animal models and humans, this reorganization includes changes in somatotopic representations and shifts in excitation/inhibition balance [2–5]. While these plastic changes may be partly adaptive, the frequent persistence of complications such as neuropathic pain and spasticity suggest that plasticity is not always well regulated. For example, SCI individuals with neuropathic pain exhibit altered corticomotor excitability and gamma-aminobutyric acid (GABA)-ergic inhibitory activity compared to those without neuropathic pain [6]. These alterations in excitability may point towards an impairment in mechanisms that normally stabilise neural activity. One such mechanism is homeostatic plasticity: the brain’s ability to maintain neuronal excitability within optimal bounds despite ongoing plastic changes [7, 8]. If these stabilising processes are compromised, shifts in excitability could fuel maladaptive reorganisation and persistent symptoms such as spasticity and neuropathic pain [9, 10].

In able-bodied individuals, homeostatic plasticity can be probed using non-invasive brain stimulation paradigms [11]. These paradigms apply an initial block of “priming” stimulation to temporarily shift corticomotor excitability, followed by a second block of “test” stimulation that reveals how this prior activity influences the brain’s plasticity. For example, delivering two blocks of anodal transcranial direct current stimulation (tDCS) in close succession typically reverses the expected outcome of the second stimulation, from facilitation to suppression [12]. This suppression is taken as an empirical indication of homeostatic plasticity, and has been shown to be reproducible and reliable in other investigations [10, 13]. The emergence of homeostatic effects depends on several parameters including the duration of stimulation, polarity, and the inter-stimulation interval. Short gaps (e.g., 3–10 minutes) tend to preserve priming effects and elicit homeostatic interactions, whereas longer gaps (≥20–30 minutes) attenuate them [14, 15].

Non-invasive brain stimulation studies using test-priming paradigms suggest that certain neurological and psychiatric populations do not show the expected homeostatic response. Specifically, following repeated excitatory stimulation, individuals with cognitive decline [16], Parkinson’s disease [17], chronic pain [9] and focal hand dystonia [18] have shown reduced suppression or even facilitation of corticomotor excitability. This lack of suppression has been proposed to reflect a disruption in homeostatic plasticity mechanisms, which normally help stabilise excitability and maintain adaptive sensory, motor, and cognitive function. It is therefore plausible that homeostatic plasticity mechanisms may also be impaired following SCI, which remains to be investigated. This is a critical question because persistent complications after SCI, including neuropathic pain, may be associated with a failure of homeostatic regulation within corticomotor and broader sensorimotor networks.

Accordingly, this study examines whether individuals with SCI retain the capacity to suppress corticomotor excitability in response to a repeated anodal tDCS protocol. The primary comparison was the difference in corticomotor excitability between individuals with SCI and able-bodied controls when anodal tDCS is preceded by anodal priming. Additional aims were to compare the effects of anodal tDCS when preceded by cathodal priming (polarity control) and sham priming (excitatory control). We hypothesised that the SCI group would show reduced homeostatic suppression of corticospinal excitability in response to repeated anodal tDCS, compared with able-bodied controls. In contrast, we expected controls to show intact homeostatic modulation, evidenced by attenuated facilitation or net suppression following two blocks of anodal tDCS (relative to baseline).

## Methods

### Participants

Twenty individuals with a SCI and 20 able-bodied controls were enrolled. Participants with SCI were eligible if they 1) had a thoracic or below SCI, 2) were aged ≥18 years, 3) their SCI occurred more than 6 months prior to their inclusion, and 4) had normal hand function. SCI participants were excluded if they 1) could not read or understand English, or 2) were contraindicated for transcranial magnetic stimulation (TMS) or tDCS according to guidelines [19], including a personal or first-degree family history of seizures, any history of stroke, serious head injury/illness resulting in brain damage, metal inside the head (e.g., surgical clips or metal fragments), implanted devices (e.g., cochlear implants, pacemakers), or pregnancy/lactation. Able-bodied controls were eligible if they were 1) aged ≥18 years and were excluded if they 1) could not read or understand English, or 2) met any TMS/tDCS contraindication listed above. Individuals with SCI were approached at arm’s length via the Neuroscience Research Australia (NeuRA, Randwick, NSW, Australia) and the NeuroRecovery Research Hub (UNSW Sydney, NSW, Australia) databases of individuals who had previously consented to be contacted about research. Able-bodied volunteers were recruited via notices at UNSW and NeuRA and general community flyers. The study received UNSW Human Research Ethics Committee approval (irecs7215). Written informed consent was obtained before any study procedures.

### Sample size and power

The primary estimand was the between-group difference (SCI *vs* controls) in the mean normalised MEP amplitude over the first 30 minutes (averaged across the 0, 5, 10, 15, 20, 25, and 30-minute timepoints) after two blocks of anodal tDCS. This 0-to-30-minute interval was selected because the homeostatic response following two blocks of anodal tDCS is typically strongest during this period [12, 13, 15]. We assumed a two-sided α=0.05 and 80% power, and targeted a standardized effect-size range of Cohen’s d = 1.05–1.46 for the primary between-group contrast. This was informed by a prior study of homeostatic plasticity in which individuals with and without chronic pain were compared on the same outcome: the mean MEP change over the 0–30 min period following two blocks of anodal tDCS (assessed at 10-min intervals) [10]. Under these assumptions, a two-sample t-test approximation indicated that n = 9-16 participants per group would detect d=1.05-1.46. We recruited n = 20 per group (total N = 40) to buffer attrition and unusable datasets, maintain ≥80% power for the primary between-group contrast, and support secondary analyses, including stratified comparisons (e.g., neuropathic pain vs no neuropathic pain). Multiplicity adjustment for the broader set of confirmatory contrasts was applied at the analysis stage and was not used as the basis for the initial sample size calculation.

### Study Protocol

All interested participants were screened for eligibility via telephone. During screening, all participants completed the TMS safety screening [19], while participants with SCI additionally provided details about injury characteristics and comorbidity (detailed in the next section). Eligible individuals were scheduled for three laboratory visits, each separated by at least fourteen days for washout. Prior to the first session, all participants completed the Beck Depression Inventory-II [20] and the State–Trait Anxiety Inventory [21] via Qualitrics. Data collection was performed at NeuRA.

The three session conditions were: (i) anodal-anodal, (ii) cathodal-anodal, and (iii) sham-anodal, with the order randomly determined per participant. Participants were blinded to session condition. Each session consisted of two 10-minute tDCS blocks separated by a 5-minute interval, with the second block always being anodal. This 10–5–10-minute design was chosen to meet the timing requirements for inducing homeostatic metaplasticity. Short inter-block intervals (3 min and 10 min) are critical for preserving the after-effects of the priming stimulation and enabling metaplastic interactions [15]. Specifically, delivering two excitatory blocks with a short pause tends to reverse the effect of the second block, producing suppression compared to a single block, whereas longer pauses (≥20–30 minutes) abolish the priming influence. A 5-minute interval therefore maintains the primed state from the first anodal stimulation, allowing us to test for homeostatic regulation. In addition, our 10–5–10 protocol aligns with prior studies showing effective cathodal priming when both priming and test blocks were 10 minutes long. These studies demonstrated enhanced plasticity or motor performance following cathodal–anodal stimulation using similar timing [14, 22, 23]. Thus, this design strikes a practical balance: it falls within the homeostatic plasticity window identified by Fricke et al. [15] while also supporting investigation of both anodal and cathodal priming effects. Corticomotor excitability (motor evoked potentials [MEPs] to transcranial magnetic stimulation) was sampled at baseline prior to priming tDCS, immediately after priming tDCS and then post-test tDCS immediately up to 60 minutes in 5-minute steps. Each MEP assessment took 2 minutes. Figure 1 shows a diagram of the protocol.

**Figure 1.**
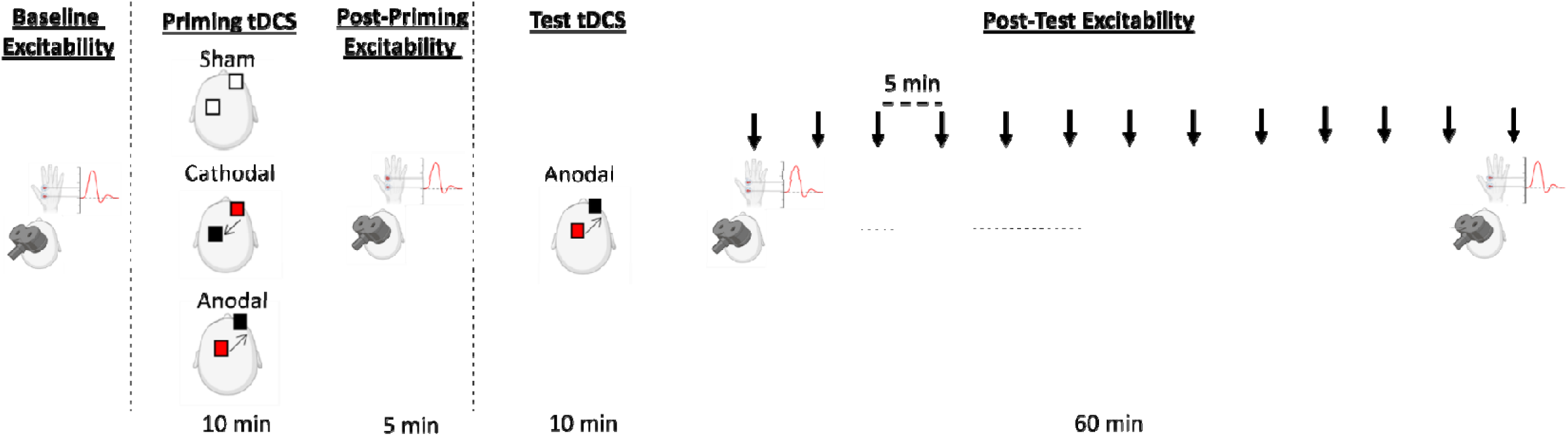
Experimental protocol. Baseline MEP amplitude was assessed, followed by 10 min of priming transcranial direct current stimulation, delivered as either anodal, cathodal or sham stimulation targeting M1. After a 5 min interval, MEP amplitude was reassessed. Test stimulation was then delivered and was always anodal transcranial direct current stimulation. MEP amplitude was subsequently assessed every 5 min for 60 min..

### Data Collection Procedures

#### SCI injury characteristics (recorded for the SCI group only)

Participants provided key injury details: neurological level (e.g., thoracic), whether injury was complete or incomplete, time since injury, source of injury, medications (type, purpose and dosage) and presence of neuropathic pain (using the 4-item SCIPI [24]). The presence of neuropathic pain was confirmed if they scored ≥2 on the 4-item SCIPI [24].

#### Beck Depression Inventory-II

A 21-item self-report measure of depressive symptoms over the past two weeks. Each item is scored 0–3; totals range 0–63, with higher scores indicating more severe symptoms [20]

#### State–Trait Anxiety Inventory

A widely used anxiety measure with two 20-item scales: State (how anxious the person feels “right now”) and Trait (how they generally feel). Items are rated on a 4-point scale; each subscale yields a score from 20–80, where higher values reflect greater anxiety [21]

#### Transcranial Magnetic Stimulation (TMS) and Electromyography (EMG)

Surface EMG from right first dorsal interosseous (FDI) used Ag/AgCl electrodes in a belly–tendon montage with distal reference and forearm ground. Signals were amplified, band-pass filtered 20–1000 Hz, notch-filtered at mains frequency as needed, and sampled at ≥2000 Hz. Single-pulse TMS was delivered with a figure-of-eight coil (Magstim, UK) over left primary motor cortex. The coil lay tangential to the scalp and induced a posterior–anterior current. The hotspot for the right FDI was identified, as defined as the scalp site yielding the largest, most consistent motor-evoked potentials – this hotspot was marked using Neuronavigation software (Neural Navigator, Neurosoft, Russia) to maintain consistency in coil orientation and angle throughout the session. Resting motor threshold (RMT) was the lowest intensity evoking ≥50 µV peak-to-peak responses in ≥5 of 10 relaxed FDI trials. Test intensity was then set to elicit ∼0.5-1 mV MEPs [18, 25], and this intensity was used for the remainder of the session. Test stimulation intensity were re-determined at the beginning of each session. Each of the target corticomotor excitability assessments was conducted by delivering 30 pulses using a jittered 2-3 inter-stimulus interval (thus each assessment was approximately 2 minutes in total). During each assessment, participants remained at rest, fixed on a fixation cross (during MEP data collection) and were instructed to relax the FDI.

#### Transcranial Direct Current Stimulation

tDCS was delivered using a battery-driven, constant-current stimulator (Soterix Medical, USA) with 5 cm circular rubber electrodes fitted with saline-soaked sponge pads (approximately 20 cm²). Before stimulation, the skin was cleaned, hair was parted, and electrode locations were marked to ensure consistent placement across sessions. Electrode impedance was continuously monitored and maintained below 10 kΩ. The target electrode was centred over the left primary motor cortex (M1) at the FDI hotspot, and the reference electrode was placed over the right supraorbital area. Stimulation was delivered at 2 mA for 10 minutes per block, with a 10 second ramp-up and ramp down period. For anodal stimulation, current entered through the M1 electrode and exited through the right supraorbital electrode. For cathodal stimulation, the current direction was reversed, entering through the right supraorbital electrode and exiting through the M1 electrode. For sham stimulation, current initially entered through the M1 electrode, but after the 10 second ramp-up, it was reduced to 0 mA for the remainder of the 10-minute period, before briefly ramping up and again at the end to mimic the sensation of active stimulation.

### Data Pre-Processing

MEP data were processed using a custom MATLAB script (Mathworks Inc., Sherborn, MA, USA). For each TMS trial, the EMG trace was segmented around the stimulus and the MEP was quantified as the peak-to-peak amplitude within a predefined post-stimulus window. Specifically, the script identified the maximum and minimum EMG values between 15 and 50 ms after the pulse and computed their difference as the trial-level MEP amplitude. Trials were then grouped into timepoint bins of 30 consecutive stimuli, and MEP amplitudes were averaged within each bin to generate a time course for each stimulation session. Timepoints comprised a session-specific baseline, an immediate post-priming timepoint, and post-test measurements from 0 to 60 minutes after the intervention, sampled in 5-minute intervals

For the primary analyses, MEP amplitudes were normalised within each session by dividing each timepoint mean by the session baseline mean. The primary outcome was expressed as the percent change from this session-specific baseline, calculated by converting the normalised values to a percent change metric. Unless stated otherwise, inferential analyses focused on the post-test period (0–60 minutes).

### Statistical Analysis

All analyses were conducted using a custom script in MATLAB. Primary inference drew from frequentist statistics. In parallel, for some analyses, evidence strength was summarised using Bayes factors derived from a Bayesian information criterion approximation based on the observed test statistics. Bayes factors were expressed as BF_10_ values, where BF_10_’s of 1–3, 3–10, 10–30, 30-100 and >100 indicated ‘weak’, ‘moderate’, ‘strong’, ‘very strong’ and ‘extreme’ evidence for the alternative hypothesis, while BF_10_’s of 1/3–1, 1/10-1/3, 1/30-1/10 and 1/100-1/30 indicated ‘anecdotal’, ‘moderate’, ‘strong’, ‘very strong’ and ‘extreme’ evidence in favour of the null hypothesis [26].

#### Stimulation Intensities and Baseline MEPs

The raw (non-normalised) baseline MEP amplitudes were compared between groups using Welch’s independent-samples t-tests and Bayes Factors.

#### Effect of Priming tDCS

To characterise the immediate effect of priming, we tested whether the normalised MEP at the post-priming timepoint differed from baseline. This was evaluated using one-sample t-tests for the full sample and separately within the control and SCI groups, again reporting both p-values and Bayes factors

#### Primary Analyses

##### Main Statistical Model

Percent change from baseline values were modelled using linear mixed-effects modelling, with a random intercept for participant to account for repeated measurements. Fixed effects included group (SCI versus control), session (anodal-anodal, cathodal-anodal, sham-anodal), time (treated as a categorical factor across post-test timepoints), and all interactions among these factors. The confirmatory analyses comprised seven pre-specified contrasts derived from the primary linear mixed-effects model. These contrasts were selected a priori to test the hypothesised effects of anodal priming, cathodal priming, and sham priming on post-stimulation corticomotor excitability.

##### Contrast 1: Between-group difference following anodal priming

The primary confirmatory contrast examined the between-group difference in the anodal-anodal condition during the first 30 minutes after test stimulation. This was operationalised as an a priori planned contrast from the linear mixed-effects model, estimating the SCI minus control difference averaged across the 0–30-minute post-test bins. Model-based effect estimates were reported together with their associated uncertainty and statistical evidence.

##### Contrasts 2 and 3: Difference from baseline within each group following anodal priming

Additional pre-specified contrasts examined the response to anodal-anodal stimulation within each group separately. Specifically, we tested whether controls showed net suppression relative to baseline, consistent with homeostatic inhibition, and whether participants with SCI showed net facilitation relative to baseline, consistent with reduced or absent homeostatic regulation [10]

##### Contrasts 4 to 7: Early effects of cathodal priming and no priming

We also evaluated the early (0–30 minute) effects of the cathodal-anodal and sham-anodal conditions. Based on prior work, we hypothesised that both cathodal priming and sham priming would be associated with a net excitatory response across the first 30 minutes, without meaningful group differences [12, 27]. Accordingly, for each of these two conditions, model-based contrasts from the primary linear mixed-effects model were used to test: (i) the overall change from baseline collapsed across groups, and (ii) the between-group difference (SCI minus controls).

##### Type I error control

Because these seven contrasts were specified as a single confirmatory family, family-wise error was controlled across all seven tests using the Holm-Bonferroni procedure. This approach preserves strong control of the family-wise error rate while being less conservative than a simple Bonferroni correction

#### Exploratory Analysis

##### Additional Contrasts

As exploratory secondary analyses, we examined the effects of anodal priming, cathodal priming, and no priming during the late post-test period (35–60 minutes) and across the full post-test interval (0–60 minutes). For each condition, we evaluated the overall change, the change within each group separately, and the between-group difference.

##### Influence of priming relative to sham

Further, we quantified the difference in MEP amplitudes between sham-anodal and anodal-anodal/cathodal-anodal (averaged across the 0–30-minute time window), which allowed us to isolate the added influence of priming on excitability while controlling for the response to anodal stimulation alone. These values were compared between groups using Welch independent sample t-tests and Bayes factors. Additional analyses were performed for each group compared to zero using one sample t-tests.

##### Subgroup Analysis

We also determined whether the early anodal-anodal plasticity (0–30 minutes) differed according to clinical characteristics in the SCI group. Subgroup comparisons included neuropathic pain status, use of medications with primarily central nervous system effects (yes/no), traumatic versus non-traumatic aetiology, completeness of injury, and a median split of time since injury. These subgroup effects were tested using Welch’s independent-samples t-tests

#### Sensitivity analyses

We examined the robustness of the primary contrast (contrast 1) in two ways. First, the analysis was repeated using absolute change derived from raw MEP amplitudes rather than from normalised time courses. Second, window summaries were recomputed using medians rather than means, and the primary analysis was repeated using these median-based summaries.

## Results

### Participant Characteristics

All participants attended their 3 sessions, with no dropouts or missing data. Table 1 summarises the characteristics of the 20 participants with SCI included in the study. The SCI cohort comprised 14 males and 6 females, with a mean age of 45.5 years (SD 10.0; range 28.4–63.1). Injuries were predominantly thoracic, with rostral neurological levels ranging from T3 to L4 (18/20 thoracic; 2/20 lumbar, L1 and L4). Injury completeness was weighted toward complete injuries (12 complete; 8 incomplete). Time since injury was highly variable (mean 16.0 years, SD 14.3; range 0.7–46.4), indicating substantial heterogeneity in chronicity. Injury causes were most commonly vehicle-related (n = 7) and sports-related (n = 5), with additional injuries due to falls (n = 3) and non-traumatic/medical aetiologies (n = 5) (stroke, neuroblastoma, tumour, neuromyelitis optica, and transverse myelitis). Neuropathic pain was reported by 13 participants (65%). Medication use was common in the SCI cohort, with 16/20 participants (80%) reporting at least one regular medication and 4/20 (20%) reporting none. The most frequently reported agents were bladder anticholinergics (solifenacin or oxybutynin; 7/20, 35%), gabapentinoids for neuropathic pain/related symptoms (pregabalin or gabapentin; 8/20, 40%). Baclofen for spasticity/spasms was reported by 3/20 (15%), and bowel agents (e.g., macrogol, docusate) by 2/20 (10%). Opioid/analgesic medications were less common (tapentadol or methadone; 3/20, 15%). Antidepressants were reported by 2/20 (10%) (amitriptyline and/or sertraline), and ADHD medication (methylphenidate) by 2/20 (10%). A single participant reported corticosteroid therapy (1/20, 5%), diuretic therapy (1/20, 5%), immunosuppressive therapy (1/20, 5%), and urinary tract infection prophylaxis (1/20, 5%).

**Table 1.** Characteristics of Spinal Cord Injured Group.

| Sex | Age Range | Injury level | Rostral level | Injury completeness | Medications | Time since injury (y) | Injury cause | Neuropathic pain |
| --- | --- | --- | --- | --- | --- | --- | --- | --- |
| M | 30-40 | T5 | T5 | Complete | **Pregabalin** (neuropathic pain)<br>**Baclofen** (spasms)<br>**Solifenacin** (Vesicare, overactive bladder)<br>**multivitamins** (magnesium). | 0.7 | Sport (cycling) | Yes |
| M | 30-40 | T10, L3 | T10 | Complete | **Pregabalin** PRN (itchiness)<br>**Solifenacin** (Vesicare, overactive bladder). | 2.1 | Vehicle (motorcycle) | No |
| F | 40-50 | T12, L1, L2, L3, L4, L5 | T12 | Incomplete | None. | 40.2 | Other (non-traumatic) | Yes |
| M | 30-40 | T10 | T10 | Complete | **Gabapentin** (neuropathic pain). | 34.2 | Other (non-traumatic) | Yes |
| M | 50-60 | T9 | T9 | Incomplete | **Gabapentin** (neuropathic pain). | 10.3 | Vehicle (hit by truck) | Yes |
| M | 50-60 | T3, T4 | T3 | Complete | **Oxybutynin** (Oxytrol, overactive bladder)<br>**Tapentadol** (Palexia, shoulder muscle pain). | 4.2 | Sport (motorcycle riding) | No |
| M | 50-60 | T5 | T5 | Complete | **Oxybutynin** (Oxytrol, overactive bladder). | 32.3 | Vehicle (motorcycle) | Yes |
| M | 40-50 | L1, L2 | L1 | Incomplete | **Gabapentin** (neuropathic pain)<br>**Macrogol** (Movicol, bowel). | 12.3 | Fall (roof collapse) | Yes |
| M | 40-50 | T12 | T12 | Incomplete | None. | 20.4 | Vehicle (run over by car) | Yes |
| M | 60-70 | T8 | T8 | Complete | **Oxybutynin** (Oxytrol, overactive bladder);<br>**Amiloride/hydrochlorothiazide** (Moduretic, ankle oedema/diuretic). | 46.4 | Vehicle (motorcycle) | No |
| M | 40-50 | T7, T8 | T7 | Incomplete | **Methylphenidate** (Ritalin, ADHD). | 23.5 | Vehicle (motorcycle) | No |
| M | 50-60 | T6 | T6 | Complete | None. | 25.6 | Sport (motocross/dirt bike) | Yes |
| M | 30-40 | T12 | T12 | Incomplete | **Solifenacin** (Vesicare, overactive bladder). | 3.6 | Vehicle (motorcycle) | No |
| F | 40-50 | T11, T12, L1, L2 | T11 | Incomplete | **Pregabalin** (Lyrica, neuropathic pain);<br>**Amitriptyline** (complex regional pain)<br>**Tapentadol** (Palexia, muscle pain)<br>**Sertraline** (depression)<br>**Baclofen** (muscle relaxant) | 25.7 | Fall (from height or level ground) | Yes |
|  |  |  |  |  | **Estradiol** (Femoston, menopause). |  |  |  |
| F | 40-50 | T7, T8, T9, T10, T11, T12 | T7 | Complete | **Dexamethasone**<br>**Gabapentin** (neuropathic pain)<br>**Methadone** (pain)<br>**Amitriptyline** (depression)<br>**Sucralfate** (ulcer). | 4.8 | Other (non-traumatic) | Yes |
| M | 40-50 | L4, L5, S1-S4 | L4 | Complete | **Methylphenidate** (Ritalin, ADHD). | 3.8 | Sport | No |
| M | 20-30 | T11 | T11 | Incomplete | **None** | 5.7 | Fall (from height or level ground) | Yes |
| F | 50-60 | T5 | T5 | Complete | **Pregabalin** (Lyrica, neuropathic pain). | 2.0 | Other (non-traumatic) | Yes |
| F | 50-60 | T7 | T7 | Complete | **Solifenacin** (Vesicare, overactive bladder). | 18.0 | Sport (equestrian) | No |
| F | 40-50 | T10 | T10 | Complete | **Mycophenolate mofetil** (immunosuppressant);<br>**Hydroxychloroquine** (Plaquenil)<br>**Baclofen** (spasticity)<br>**Methenamine hippurate** (Hiprex, UTI prevention)<br>**Pregabalin** (Lyrica, neuropathic pain);<br>**Solifenacin** (Vesicare, overactive bladder);<br>**Docusate sodium** (Coloxyl, laxative);<br>**Macrogol** (Movicol, bowel). | 4.0 | Other (non-traumatic) | Yes |

Table 2 shows the mean age, sex distribution, anxiety, depression and for the SCI and control groups. The male and female split was identical. Furthermore, the Bayes and Welch independent t-tests revealed evidence of no significant differences between groups in age, *t*(33.23) = -0.289, *p* = 0.775, BF_10_ = 0.32, BDI score, *t*(37.84) = 0.062, *p* = 0.95, BF_10_ = 0.16, and STAI score, *t*(36.97) = 0.47, *p* = 0.639, BF_10_ = 0.18. Together, these results provide moderate evidence that the SCI and control samples were well matched on age, sex, and psychiatric symptom severity.

**Table 2.** Demographics of SCI and control Group.

| Group | N | Female n (%) | Male n (%) | Age, mean (SD) | STAI-State, mean (SD) | BDI, mean (SD) |
| --- | --- | --- | --- | --- | --- | --- |
| Controls | 20 | 6 (30.0%) | 14 (70.0%) | 44.0 (14.80) | 32.70 (9.15) | 5.5(7.88) |
| SCI | 20 | 6 (30.0%) | 14 (70.0%) | 45.15 (9.93) | 34.20 (10.83) | 5.65 (7.39) |

### Stimulation Characteristics

Averaged across the three sessions, RMT did not differ between groups with moderate Bayesian evidence, *t*(34.75) = 0.880, *p* = 0.385, BF*_10_* = 0.238. Across sessions, test stimulation intensities (%MSO) were not significantly different between groups, with moderate Bayesian evidence, *t*(37.2) = −0.56, *p* = 0.579, BF*_10_* = 0.19.

### Baseline corticomotor excitability

There was no significant group difference in baseline MEP amplitude for the anodal-to-anodal condition, *t*(36.0) = 1.52, *p* = 0.14, BF_10_= 0.55, cathodal-to-anodal condition, *t*(33.0) = −0.78, *p* = 0.44, BF_10_= 0.23, or in the sham-to-anodal condition, t(37.4) = 0.04, p = 0.969, BF_10_ = 0.16. This provides anecdotal to moderate support for the null hypothesis of no baseline group difference across sessions.

### Normalised priming effect

In the anodal-anodal condition, there was an overall trend towards an increase in MEP amplitude from baseline which did not reach significance, *t*(39) = 1.76, *p* = 0.086, BF_10_ = 0.73. In the cathodal-anodal condition, there was an overall trend towards a reduction from baseline which was also non-significant, *t*(39) = −1.81, *p* = 0.077, BF_10_ = 0.80. In the sham-anodal condition, there was no overall difference from baseline following sham tDCS, *t*(39) = −0.30, *p* = 0.7671, BF_10_ = 0.17

### Primary Analysis

Figure 2 shows the change from baseline in MEP amplitude for the three different conditions separated by group, across the 60-minute period post tDCS. These data were analysed using a linear mixed-effects model with fixed effects for group, session, time (categorical across post-test timepoints), and all interactions, with a random intercept for participant. The primary post-test model was fitted to 1,558 observations from 40 participants.

**Figure 2.**
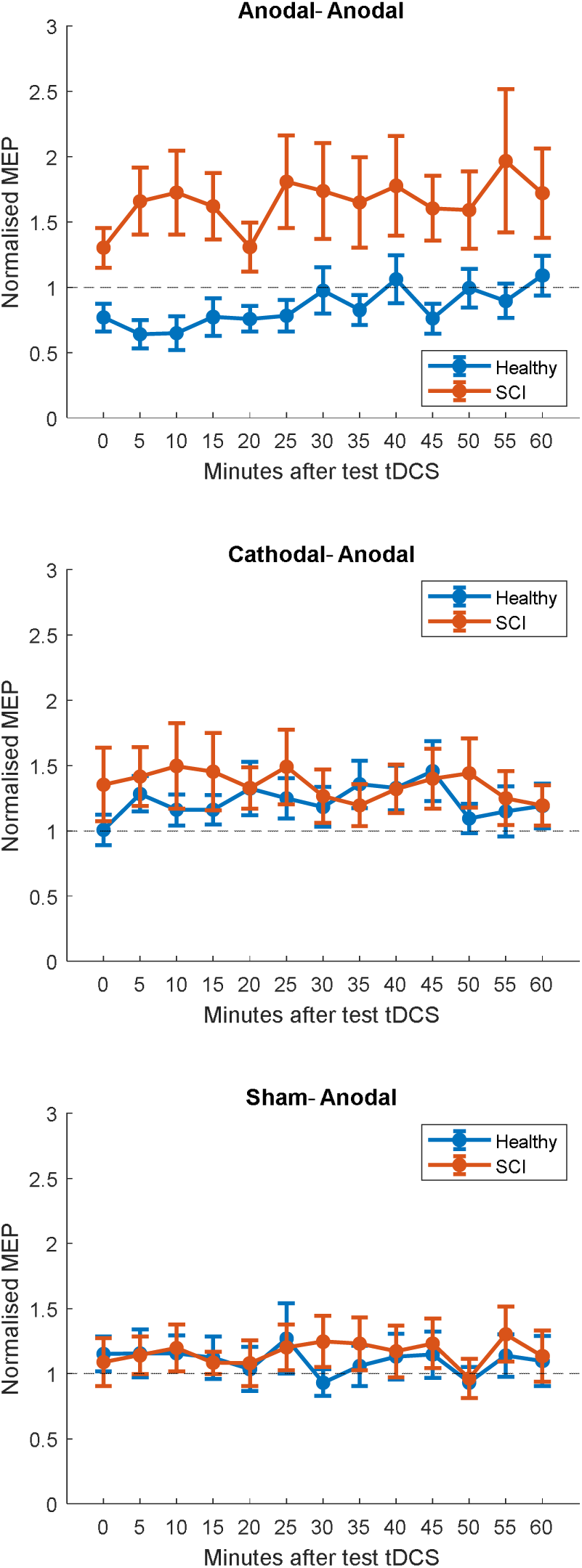
Normalised MEP amplitude at each post-test tDCS timepoint for each condition in healthy controls (blue) and the SCI group (red).

#### Contrast 1: Between-group difference following anodal priming

For the anodal-anodal condition during the first 30 minutes after test stimulation, the SCI group showed a greater mean ΔMEP% than controls, with a model-based contrast estimate of 83.09 ΔMEP% (SE = 17.00), *t*(1480) = 4.89, *p* < 0.001, 95% CI [49.75, 116.43].

#### Contrasts 2 and 3: Difference from baseline within each group following anodal priming

In the 0-30 minutes following anodal-anodal stimulation, controls showed a reduction in MEP amplitude relative to baseline (estimate = -23.62 ΔMEP%, SE = 12.02, *t*(1480) = -1.97, *p* = 0.0495, 95% CI [-47.20, -0.05]), whereas participants with SCI showed an increase relative to baseline (estimate = 59.46 ΔMEP%, SE = 12.02, *t*(1480) = 4.95, *p* <.001, 95% CI [35.89, 83.04]).

#### Contrasts 4 to 7: Early effects of cathodal priming and no priming

For the cathodal-anodal condition over the first 30 minutes, there was evidence of an overall increase from baseline when collapsed across groups (estimate = 29.79 ΔMEP%, SE = 8.50, *t*(1480) = 3.51, *p* <.001, 95% CI [13.12, 46.46]), but no evidence of a between-group difference (estimate = 20.49 ΔMEP%, SE = 17.00, *t*(1480) = 1.21, *p* = 0.23, 95% CI [-12.85, 53.83]).

For the sham-anodal condition over the first 30 minutes, there was no clear evidence of an overall change from baseline (estimate = 13.30 ΔMEP%, SE = 8.50, *t*(1480) = 1.57, *p* = 0.12, 95% CI [-3.37, 29.97]) and no evidence of a between-group difference (estimate = 3.11 ΔMEP%, SE = 17.00, *t*(1480) = 0.18, *p* = 0.86, 95% CI [-30.24, 36.45]).

After Holm -Bonferroni correction across the seven confirmatory contrasts, only three effects remained statistically significant: the primary anodal-anodal between-group contrast, the anodal-anodal SCI-within-group facilitation contrast, and the cathodal-anodal overall facilitation contrast.

### Exploratory analyses

#### Additional Contrasts (Figure 3)

In exploratory analyses, the between-group difference in the anodal-anodal condition remained evident beyond the primary early window. The SCI group continued to show greater ΔMEP% than controls in the late 35–60 minute window (estimate = 78.07 ΔMEP%, SE = 17.45, *t*(1480) = 4.48, *p* < 0.001, 95% CI [43.85, 112.30]) and across the full 0–60 minute interval (estimate = 80.77 ΔMEP%, SE = 15.68, *t*(1480) = 5.15, *p* < 0.001, 95% CI [50.01, 111.53]).

**Figure 3.**
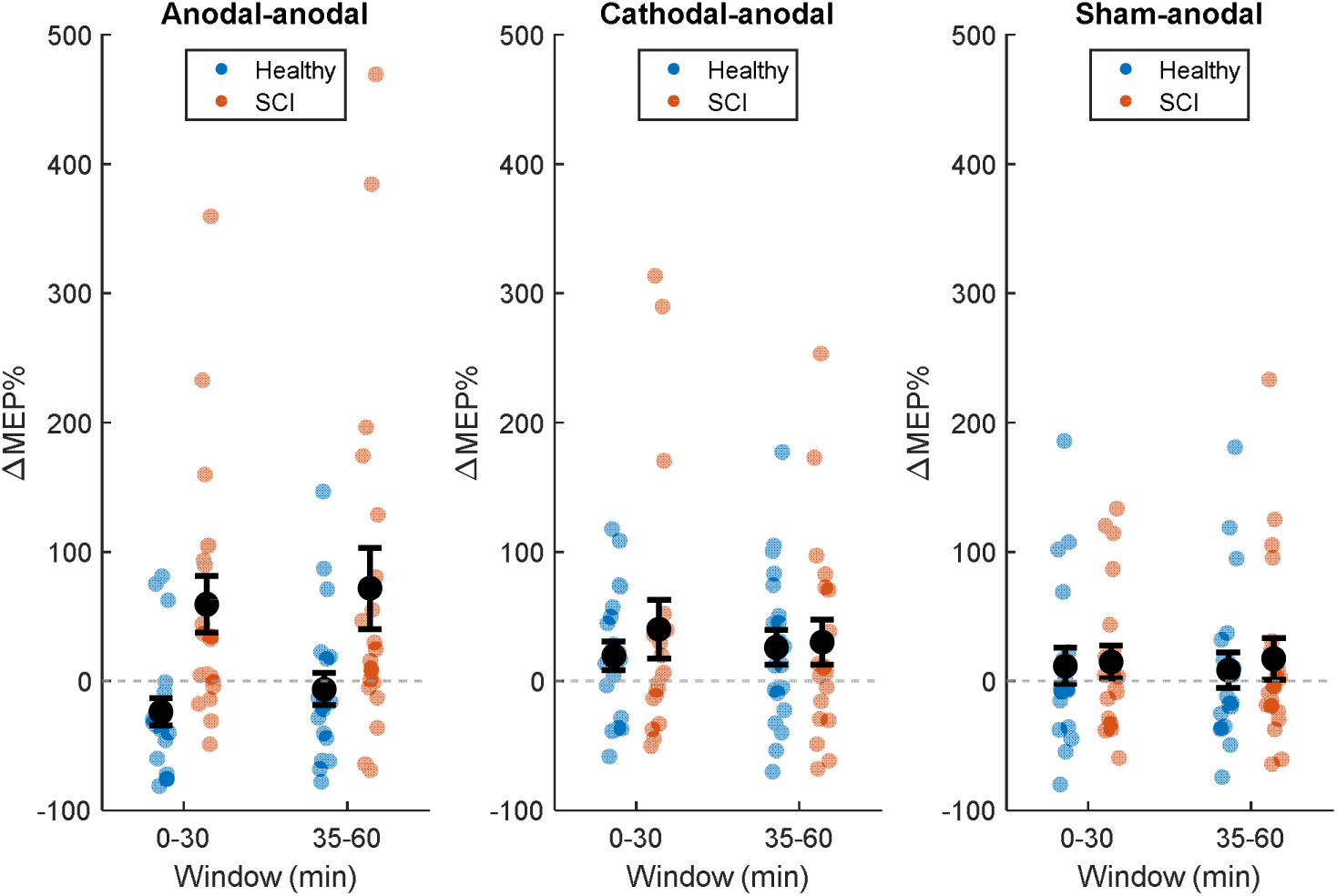
Mean (+SEM) and Individual Normalised MEP amplitudes for the 0-30 Minute and 35-60 Minute windows for each condition in healthy controls (blue) and the SCI group (red).

For the cathodal-anodal condition, collapsed across groups, ΔMEP% was increased in the late 35–60 minute window (estimate = 28.03 ΔMEP%, SE = 8.74, *t*(1480) = 3.21, *p* = 0.00137) and across the full 0–60 minute interval (estimate = 28.98 ΔMEP%, SE = 7.85, *t*(1480) = 3.69, *p* = <.0001), with no evidence of between-group differences in either window (late: *p* = 0.817; full: *p* = 0.411).

For the sham-anodal condition, there was no clear evidence of a change from baseline in the late 35–60-minute window or across the full 0–60-minute interval, whether assessed overall or as a between-group contrast (all *p* ≥ 0.095).

#### Influence of priming relative to sham (Figure 4)

When computing the effect of two blocks of anodal stimulation relative to a single block (anodal-anodal minus sham-anodal across 0-30 min timepoints) there was a significant difference between groups, with a larger value in SCI than in controls, with strong Bayesian evidence, *t*(31.6) = 2.72, *p* = 0.01, BF_10_ = 10.70. Within-group testing showed that controls differed significantly from zero in the negative direction, with anecdotal Bayesian evidence, *t*(19) = -2.30, *p* = 0.03, BF_10_ = 2.59, whereas the SCI group showed a trend in the positive direction, with anecdotal Bayesian Evidence, *t*(19) = 1.78, *p* = 0.09, BF_10_ = 1.05.

**Figure 4.**
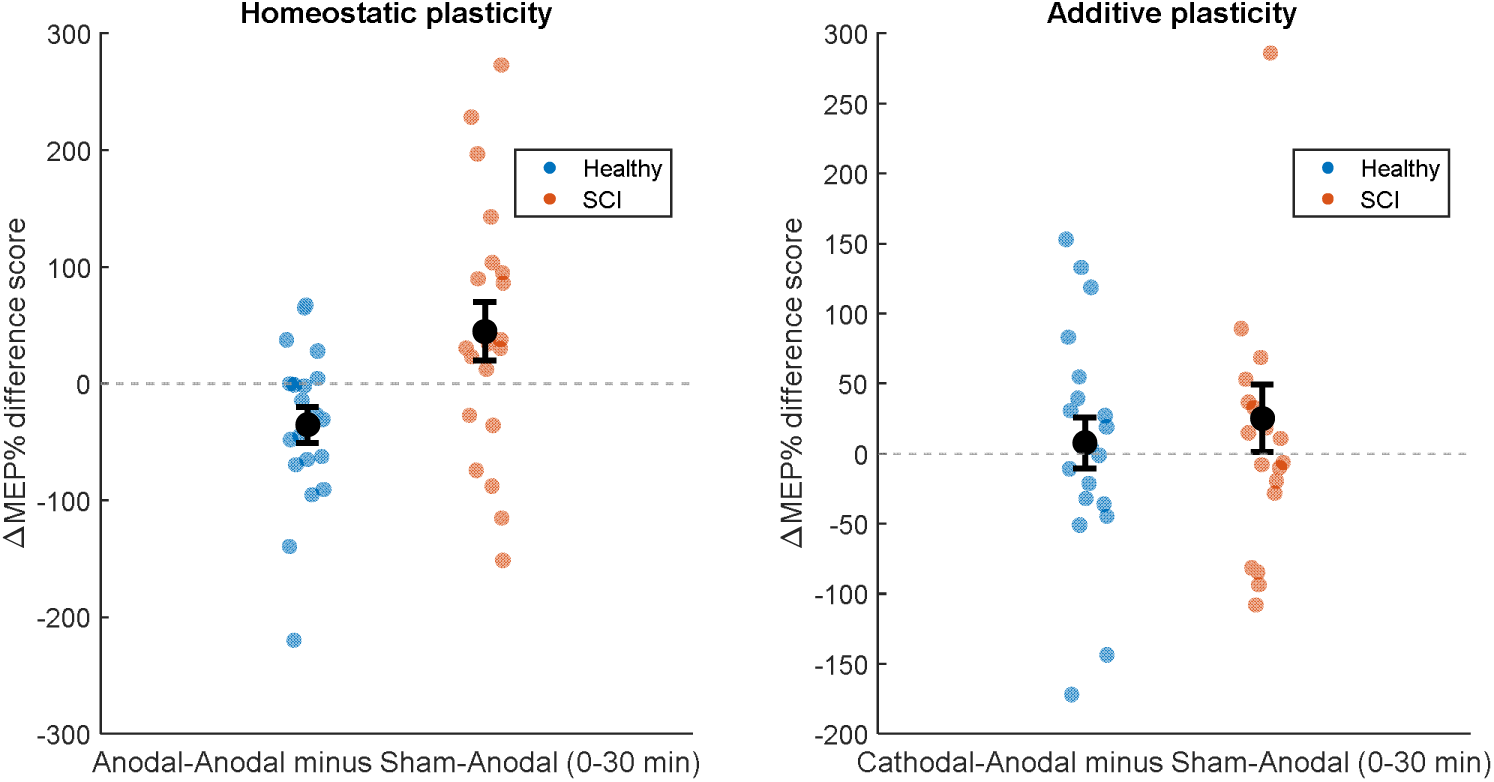
Mean (±SEM) and individual normalised MEP difference scores across the 0–30 min window for the Anodal-Anodal minus Sham-Anodal conditions (left), reflecting homeostatic plasticity, and the Cathodal-Anodal minus Sham-Anodal conditions (right), reflecting additive plasticity, in healthy controls (blue) and participants with SCI (red).

By contrast, the comparison between cathodal priming to no priming (cathodal-anodal minus sham-anodal across 0-30 min timepoints) did not differ between groups, *t*(35.4) = 0.58, *p* = 0.57, BF_10_ = 0.19, and neither controls nor SCI differed from zero (healthy: *p* = 0.67, BF_10_ = 0.25; SCI: *p* = 0.31, BF_10_ = 0.39).

##### Subgroup Analysis (Figure 5)

Within the SCI group, exploratory subgroup analyses of early anodal–anodal plasticity (0–30 minutes) suggested larger facilitation in participants with neuropathic pain than in those without neuropathic pain, *t*(17.9) = 2.203, *p* = 0.04, BF_10_ = 2.465, and in those with traumatic versus non-traumatic injuries, *t*(16.2) = 2.462, *p* = 0.025, BF_10_ = 5.34. By contrast, there was no clear evidence that early anodal–anodal responses differed by CNS-acting medication status, injury completeness, or time-since-injury group (CNS-acting medication: *t*(10.7) = −1.02, *p* = 0.33, BF_10_ = 0.58; completeness: *t*(16.2) = −0.652, *p* = 0.52, BF_10_= 0.290; time since injury: *t*(10.7) = −1.51, *p* = 0.16, BF_10_ = 1.53).

**Figure 5.**
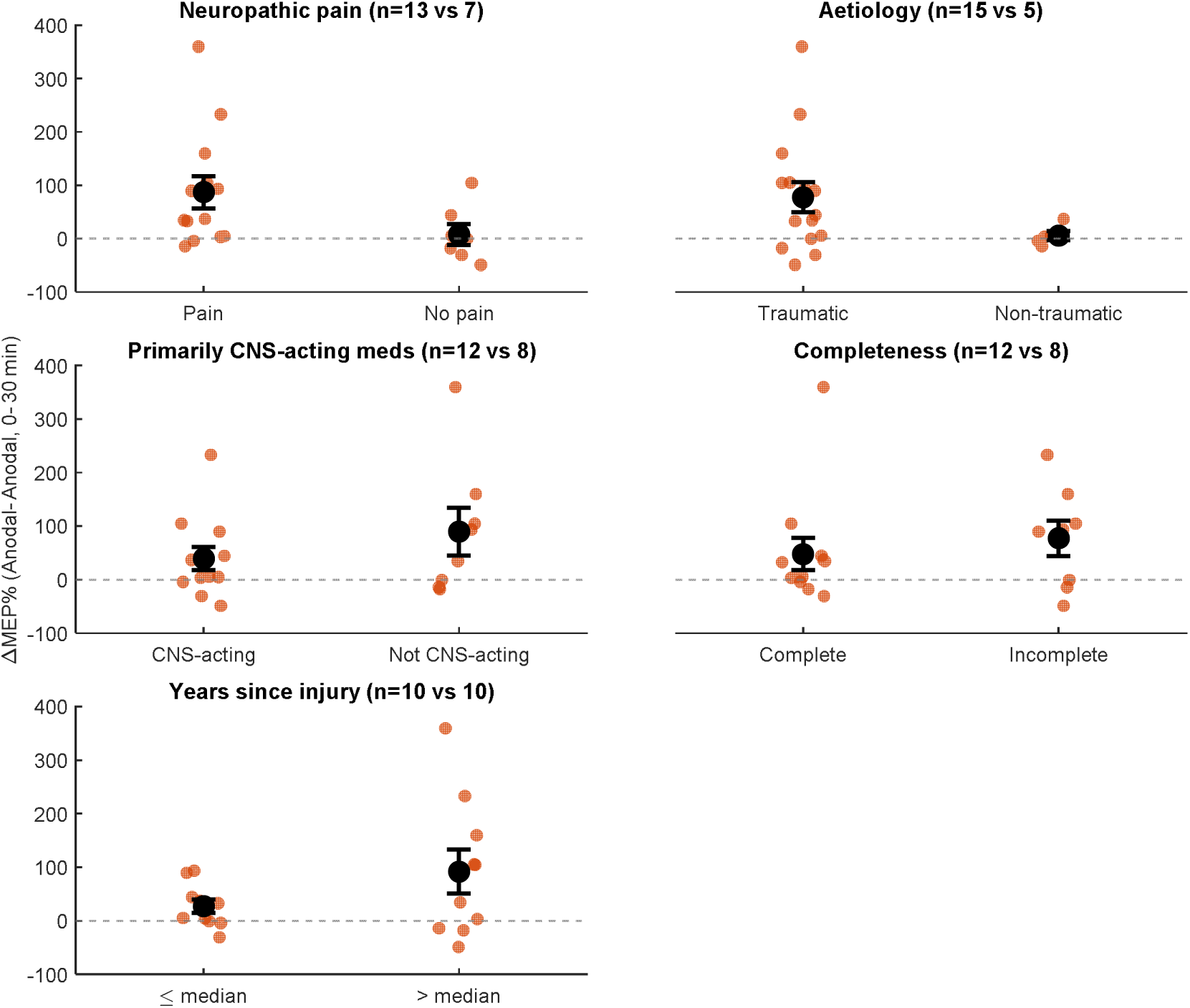
Mean (±SEM) and individual normalised MEP across the 0–30 min window for the Anodal-Anodal session for SCIs split by neuropathic pain status, injury aetiology, medication classification, years since injury (median split)

### Sensitivity analyses

The primary inference was robust across two sensitivity analyses. Fist, when the primary early anodal-anodal comparison was repeated using absolute raw MEP change (µV) rather than normalised change scores, the between-group effect remained significant, with strong Bayesian evidence, *t*(36.8) = 3.42, *p* = 0.0016, BF_10_ = 39.30. Second, when the primary comparison was repeated using median-based rather than mean-based early window summaries, the result was again unchanged, with a significant SCI minus healthy difference, with strong Bayesian evidence, *t*(30.1) = 3.53, *p* = 0.001, BF_10_ = 158.80.

## Discussion

The present study indicates that individuals with SCI show reduced homeostatic regulation of excitability to repeated excitatory stimulation. Previous studies have shown that, in healthy individuals, delivering two closely spaced blocks of anodal tDCS leads to a reduction in corticospinal excitability, which is taken as an empirical marker of homeostatic plasticity [7–10]. Our key result was a clear difference between groups following two anodal tDCS blocks (the anodal-anodal condition). Individuals with SCI showed a substantially larger increase in corticomotor excitability than healthy controls. Importantly, this between-group effect was robust: the same conclusion held when the outcome was analysed on the raw MEP scale, and when early-window summaries were computed using medians rather than means. Together, these checks suggest the finding reflects a stable physiological pattern rather than an artefact of a particular analysis choice. We also found that, when anodal stimulation was preceded by cathodal priming (the cathodal-anodal condition), excitability increased overall when collapsed across groups, with no evidence of a between-group difference. Finally, the groups were well matched on age, sex distribution, anxiety, depressive symptoms, baseline MEP amplitudes and stimulation intensities, strengthening confidence, that the observed effects are not attributable to baseline sample differences.

### Homeostatic Plasticity is Impaired following SCI

SCI is known to trigger widespread sensorimotor reorganisation, including changes in cortical representations and shifts in excitation–inhibition balance [1–5, 28]. While some reorganisation may be adaptive, persistent complications such as neuropathic pain and spasticity suggest that plasticity after SCI may not always be well regulated. Homeostatic plasticity is one of the brain’s key stabilising processes, helping maintain excitability within functional bounds despite ongoing synaptic change [8, 11]. If this regulatory system is weakened, increases in excitability may not be sufficiently counterbalanced, allowing maladaptive physiological states to emerge or persist [9, 10]. The present findings are consistent with this idea. Instead of showing the expected suppressive response after repeated excitatory input, the SCI group showed clear facilitation, whereas healthy controls showed a reduction in excitability in the early window. Although the healthy within-group suppression effect did not remain statistically significant after multiplicity correction, its direction aligns with prior work on homeostatic effects in repeated anodal stimulation paradigms [12]. Comparing the repeated-stimulation condition with the single-stimulation control further strengthened the interpretation of altered regulation after SCI. Specifically, when a homeostatic index is quantified as the difference between the anodal-anodal and sham-anodal conditions, the groups diverged: controls showed a negative shift, whereas the SCI group showed a positive shift. This contrast is informative because it captures the added impact of excitatory priming while partially controlling for the response to the anodal test block itself. Finally, the between-group difference was not confined to the early post-test period. Participants with SCI continued to show greater facilitation than controls in the late 35–60-minute window and across the full 0–60-minute interval, whereas controls tended to drift back toward baseline after the initial suppression. This temporal profile suggests that SCI may be characterised not only by an exaggerated early excitability response to repeated excitation, but also by a diminished capacity to normalise excitability back toward baseline once it has been perturbed.

### Cathodal Priming leads to net facilitatory effects

Findings from the cathodal-anodal condition provide an important complementary perspective. In the early 0-to-30-minute window, cathodal priming followed by anodal stimulation produced a significant overall increase in excitability when collapsed across groups, with this effect surviving multiplicity correction. This is broadly consistent with metaplastic models in which inhibitory priming enhances the facilitatory effect of subsequent excitatory stimulation [12, 29]. Similar facilitation was also evident in the 35-60 minute window and full 0-60 minute window. Crucially, however, there was no evidence that the cathodal-anodal effect differed between groups. This suggests that the capacity for enhancement of excitability following inhibitory priming is relatively preserved in SCI. In other words, the present results do not imply a breakdown of all metaplastic mechanisms. Rather, they point to a more selective abnormality, specifically in the suppressive homeostatic response to repeated excitation.

While in the expected direction the sham-anodal condition did not show clear evidence of an overall increase from baseline in the confirmatory 0–30 minute window, nor was there evidence of a between-group difference. This pattern is unsurprising given that anodal tDCS effects on corticospinal excitability are well known to be highly variable across individuals, and many studies do not reliably observe facilitation at the group level [30]. Importantly, one determinant of this variability is the prevailing neurophysiological “state” at the time of stimulation [31]. By using priming paradigms to manipulate state, our data indicate that the most informative effects are those reflecting state-dependent modulation of the response to anodal stimulation, rather than a simple main effect of anodal tDCS delivered in isolation.

### Impaired Homeostatic Plasticity may be associated with Neuropathic Pain and Traumatic Injury

The exploratory subgroup analyses provide an initial indication that these physiological abnormalities may be clinically meaningful. Within the SCI group, participants with neuropathic pain showed larger increases in MEP amplitude than those without neuropathic pain following two blocks of anodal tDCS. Indeed, inspection of Figure 5 suggests that those without neuropathic pain did not appear to show an increase in MEPs relative to baseline. This result compliments prior evidence that SCI individuals with neuropathic pain show altered corticospinal excitability and inhibitory dysfunction relative to those without neuropathic pain [6] such that it is not just excitability that is different but the ability to regulate that excitability. It also fits the broader proposal that impaired homeostatic regulation may contribute to maladaptive cortical states that help sustain persistent symptoms [9, 10]. In addition, early anodal–anodal plasticity differed by injury aetiology, with greater facilitation in traumatic compared with non-traumatic SCI. This suggests that the integrity of stabilising mechanisms may also vary with injury mechanism and the degree of abrupt sensorimotor deafferentation it produces [32]. By contrast, there was no clear evidence that early responses differed by CNS-acting medication status, injury completeness, or time since injury. These subgroup findings should be interpreted cautiously given limited subgroup sizes and clinical heterogeneity, and require replication in larger, more finely phenotyped cohorts.

### Clinical Implications

Clinically, these findings suggest that impaired homeostatic plasticity may be relevant not only to the pathophysiology of SCI, but also to the future development of physiology-guided assessment and treatment. A major translational gap in SCI is the absence of validated physiological biomarkers that can identify who will develop persistent complications such as neuropathic pain [33, 34], who is most likely to respond to treatment, or the neurophysiological mechanisms underlying interventions such as brain stimulation [35, 36]. Recent work has highlighted that, for SCI neuropathic pain, there is still no approved physiologically relevant biomarker, while in motor recovery, there is no simple marker that reliably identifies which patients have recovery potential [37]. Because homeostatic paradigms directly probe the capacity of the cortex to regulate excitability, the anodal-anodal response profile may provide a mechanistically meaningful marker of plasticity regulation, rather than a static index of impairment alone. This interpretation is strengthened by pain literature showing that homeostatic responses are disrupted in painful states and may recover when pain resolves, consistent with the possibility that these measures track maladaptive physiology over time [9, 10].

From a treatment perspective, the preserved facilitatory response in the cathodal-anodal condition suggests a practical opportunity for intervention design. Our findings indicate that the sequence and physiological context of stimulation may matter, with inhibitory priming potentially offering a way to shape subsequent excitatory effects in a more predictable manner, whereas excitatory stimulation alone may produce more variable responses. Indeed, although inhibitory “preconditioning” stimulation has been shown to enhance subsequent plasticity, [14, 22, 23, 29] it has rarely been explored as a therapeutic strategy.

### Limitations and future directions

Several limitations should be acknowledged. First, although the study was adequately powered for the primary anodal-anodal between-group contrast, it was not powered for subgroup analyses. In particular, the SCI sample was also clinically heterogeneous, with substantial variation in injury chronicity, completeness, neuropathic pain status, and medication use. While this improves ecological validity, it also introduces variance that may obscure more specific mechanistic patterns. Future studies should therefore seek to replicate these findings in larger and more clinically stratified SCI cohorts, with more targeted examination of neuropathic pain, spasticity, and medication effects. Second, the study focused on thoracic-and-below SCI in individuals with preserved hand function, and the findings may not generalise to cervical injuries, more severe upper-limb impairment, or other motor representations. Third, while the present findings are consistent with impaired homeostatic plasticity, they do not identify the underlying mechanism. Several candidates are biologically plausible. Abnormal N-methyl-D-aspartate (NMDA)-dependent metaplasticity is a plausible contributor, because homeostatic responses to repeated non-invasive brain stimulation are thought to depend in part on NMDA-receptor mediated plasticity mechanisms [38]. Disrupted sensorimotor integration and thalamocortical reorganisation may also be relevant, as SCI-related neuropathic pain has been linked to altered thalamic GABA, blood flow, and corticothalamic-corticospinal network activity[39]. Future studies combining test-priming paradigms with paired-pulse TMS indices of cortical inhibition, sensory pathway measures, and multimodal imaging may therefore be especially valuable for clarifying the mechanisms underlying the present effects [40]

### Conclusions

In summary, this study provides the first direct evidence that the expected homeostatic suppressive response to repeated excitatory stimulation is impaired after SCI. In contrast, facilitation following cathodal priming appeared relatively preserved, suggesting that not all forms of metaplasticity are disrupted equally. Together, these findings support the idea that persistent complications after SCI may reflect not merely the presence of plasticity, but a reduced ability to regulate it appropriately. This distinction may be important for understanding maladaptive reorganisation after SCI and for refining future neuromodulation-based treatment approaches.

## Data Availability

All data produced in the present study are available upon reasonable request to the authors

## Conflicts of Interests

None

## Funding Statement

The study was supported by the Doris and June Cather Postdoctoral Fellowship awarded to NC and the Rebecca Cooper Fellowship awarded to SG

## Notes

### Competing Interest Statement

The authors have declared no competing interest.

### Author Declarations

The study received UNSW Human Research Ethics Committee approval (irecs7215). Written informed consent was obtained before any study procedures.

